# Genomic foundation models extend clinicopathologic and transcriptomic prognostication in soft tissue sarcoma

**DOI:** 10.64898/2026.09.21.26363568

**Authors:** Miguel Esperança-Martins, Manuel Sokolov Ravasqueira, Sérgio Dias, Nuno Abecasis, Luís Costa, Hugo Vasques

**Affiliations:** Medical Oncology Department, Unidade Local de Saúde de Santa Maria, Lisboa, Portugal; Gulbenkian Institute for Molecular Medicine - CARE, Lisboa, Portugal; Faculdade de Medicina da Universidade de Lisboa, Lisboa, Portugal; Instituto Superior Técnico, Lisboa, Portugal; Instituto de Engenharia de Sistemas e Computadores - Investigação e Desenvolvimento, Lisboa, Portugal; General Surgery Department, Instituto Português de Oncologia de Lisboa Francisco Gentil, Lisboa, Portugal

**Keywords:** soft tissue sarcoma, prognosis, genomic foundation model, Evo2, SARCULATOR, CINSARC

## Abstract

**Background:** Clinicopathologic (SARCULATOR) and transcriptomic (CINSARC) models are established prognostication tools in soft tissue sarcoma, but neither directly represent the sequence context of tumor genomic alterations. Evo2 is a genomic foundation model trained to learn the grammar of DNA/RNA sequences. We tested whether Evo2-derived features from tumor short variants, fusions, and rearrangements improve survival prognostication beyond clinicopathologic and transcriptomic risk models.

**Methods:** We analysed formalin-fixed, paraffin-embedded tumor material and clinical data from 102 patients with dedifferentiated liposarcoma (n = 25), high-grade leiomyosarcoma (n = 25), and undifferentiated pleomorphic sarcoma (n = 52). A separate TCGA-SARC cohort of comparable histologies (n = 129) served as external validation. Reconstructable alterations (short variants, fusions and rearrangements) were converted into sequence inputs, embedded with Evo2 and aggregated into patient-level genomic representations. Cox proportional hazards models incorporated genomic features alone (Evo2) or together with clinicopathologic (SARCULATOR) or transcriptomic (CINSARC) predictors across overall survival (OS), progression-free survival (PFS), disease-free survival (DFS), relapse-free survival (RFS), and metastasis-free survival (MFS). Discrimination was assessed using Harrell’s concordance index (C-index).

**Results:** Evo2 features were generated from 174 study-cohort sequences and 7298 TCGA-SARC sequences. In the study cohort, adding Evo2 to SARCULATOR increased the reported C-index for OS from 0.620 to 0.770, PFS from 0.625 to 0.755, DFS from 0.612 to 0.714, and RFS from 0.633 to 0.771. In TCGA-SARC, SARCULATOR plus Evo2 increased the OS C-index from 0.595 to 0.716 and the MFS C-index from 0.548 to 0.667. Also in TCGA-SARC, adding Evo2 to CINSARC increased the OS C-index from 0.476 to 0.653 and the MFS C-index from 0.553 to 0.650. These are descriptive point-estimate comparisons. Moreover, in TCGA-SARC Evo2 alone outperformed SARCULATOR alone and CINSARC alone, supporting independent prognostic information within altered sequence-context representations.

**Conclusions:** Evo2-derived genomic representations improved clinicopathologic and transcriptomic prognostication across sarcoma cohorts and survival endpoints. These findings support genomic foundation models as an independent and complementary layer for sarcoma risk stratification through direct modeling of altered tumor sequence biology. Confirmation of model transportability, calibration and clinical utility is required before clinical application.

**Graphical abstract:** 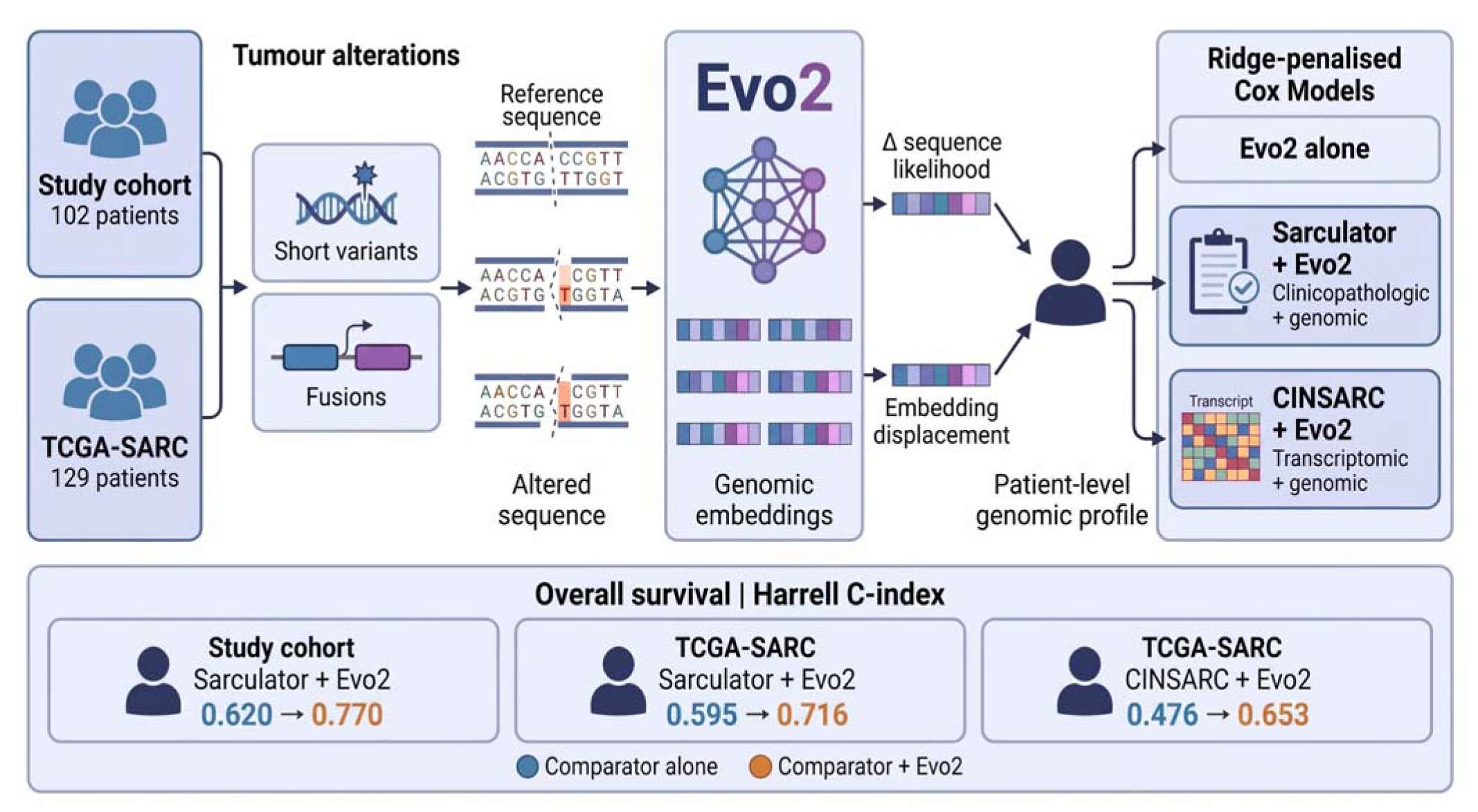

*Genomic sequence representations for sarcoma prognostication.:* Reconstructed short variants and study-cohort fusion proxies are processed with Evo2 and aggregated into patient-level genomic profiles for ridge-penalised Cox survival modelling. Genomic profiles are evaluated alone or with clinicopathologic SARCULATOR or transcriptomic CINSARC predictors; CINSARC is evaluated only in TCGA-SARC. The lower panel shows reported overall-survival Harrell C-index point estimates before and after adding Evo2 features. These descriptive comparisons do not establish statistical significance or clinical utility. Δ denotes the alternative-minus-reference difference in mean sequence log-likelihood. Embedding displacement denotes cosine distance between paired short-variant embeddings. FFPE, formalin-fixed, paraffin-embedded; TCGA-SARC, The Cancer Genome Atlas sarcoma cohort. Created using BioRender.

## Introduction

Accurate prognostication is central to managing soft tissue sarcomas (STS), whose heterogeneous biological behavior is intrinsically linked with miscellaneous patterns of recurrence, metastasis and survival. In localized and advanced disease, risk assessment is central to support decision regarding both the employment and also the timing and intensity of different treatment modalities, as well as the use of particular surveillance strategies, always balancing potential benefit against treatment morbidity [1,2]. Better individual risk estimation could increase the capacity of identifying the most suitable timing of initiation of different treatment modalities, reduce both undertreatment of aggressive tumors and unnecessary treatment of lower-risk disease, and refine the design of specific surveillance strategies.

Current prognostication approaches integrate clinicopathologic nomograms and molecular signatures. SARCULATOR provides externally validated, accessible estimates using clinicopathologic variables including age, tumor size, grade, histopathological subtype, multifocality, or completeness of resection [3]. Nevertheless, these variables are indirect biological proxies and cannot fully resolve heterogeneity within apparently similar tumors. CINSARC adds a 67-gene expression signature reflecting mitosis and chromosomal integrity, with validated associations with metastasis-free survival (MFS) [4]. Its predefined biological focus, however, does not comprehensively represent the immune, DNA damage repair and other biological regulatory processes shaping tumor behavior. Population differences, frequent histopathological subtype discordances, grading variability and assay dependence threaten transportability; CINSARC performance also varies with treatment setting [5,6]. These limitations motivate integration of complementary molecular information, provided that additional complexity yields reproducible improvement beyond established models.

In our 2025 study, unsupervised machine learning transcriptomic analysis identified four STS subtypes spanning across conventional histological categories, characterized by reduced DNA-repair gene expression, cancer-testis antigen enrichment, immune activation and claudin-related programmes [6]. Their prognostic associations were supported in independent cohorts, and their molecular profiles suggested potential specific and tailored therapeutic strategies. This past work demonstrated the value of examining biological diversity beyond morphology, while leaving open whether genomic sequence information could further refine prognostic stratification.

Genomic foundation models learn reusable numerical representations of DNA through self-supervised training on large sequence collections. Evo2 is an autoregressive model based on the StripedHyena 2 architecture, combining convolutions and attention to learn nucleotide dependencies across multiple scales [7]. Evo2 learns by predicting the next nucleotide, producing embeddings that can subsequently be used in task-specific models. Trained on genomes across domains of life, it captures features associated with evolutionary constraint, coding organization and regulatory sequences. Applied to altered tumor sequences, these representations could add contextual information about genomic perturbations beyond clinicopathologic phenotypes or measured transcript abundance. This motivates testing their prognostic performance both independently and alongside SARCULATOR and CINSARC. Indeed, sequence-level functional prediction alone does not establish survival prediction.

We hypothesized that Evo2-derived representations of tumor genomic alterations contain prognostic information complementary to established clinicopathologic and transcriptomic models. We reconstructed sequences from short variants, fusions and rearrangements, extracted Evo2 features and aggregated them into patient-level profiles in a 102-patient study cohort and in a 129-patient TCGA-SARC external validation cohort. Using Cox survival models, we evaluated genomic representations alone and their incremental prognostic discrimination when combined with SARCULATOR or CINSARC. The overall design is shown in Figure 1.

**Figure 1.**
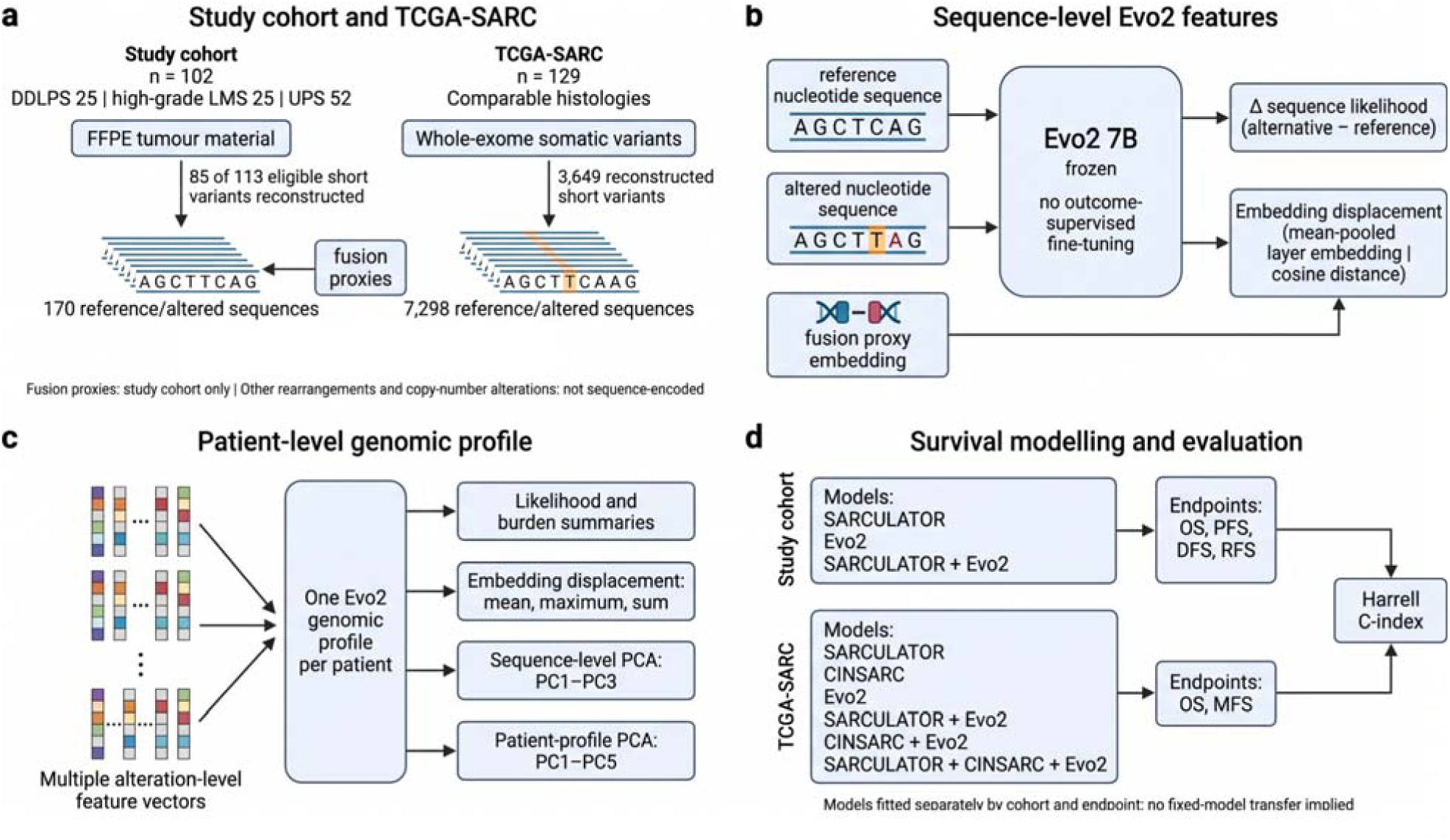
Study design and derivation of patient-level Evo2 genomic profiles. **a.** Cohort-specific sequence reconstruction. In the study cohort, 85 reconstructed short variants yielded 170 reference and alternative sequences; fusion proxies brought the total to 174 sequence inputs. TCGA-SARC contributed 3,649 reconstructed short variants and 7,298 sequences, **a**. separate cohort total. Other rearrangements and copy-number alterations were not sequence-encoded. **b**. Frozen Evo2 7B generated sequence log-likelihoods and mean-pooled embeddings. For short variants, Δ denotes alternative-minus-reference mean log-likelihood, and embedding displacement denotes cosine distance between paired embeddings. Fusion proxies contributed embeddings in the study cohort only. **c**. Patient-level profiles combined likelihood and burden summaries, embedding-displacement summaries, sequence-level principal components PC1–PC3 and patient-profile components PC1–PC5. **d**. Model families and survival endpoints evaluated using Harrell’s concordance index (C-index). Models were fitted separately by cohort and endpoint; no transfer of a fixed fitted model between cohorts is implied. DDLPS, dedifferentiated liposarcoma; LMS, leiomyosarcoma; UPS, undifferentiated pleomorphic sarcoma; FFPE, formalin-fixed, paraffin-embedded; TCGA-SARC, The Cancer Genome Atlas sarcoma cohort; PCA, principal component analysis; PC, principal component; OS, overall survival; PFS, progression-free survival; DFS, disease-free survival; RFS, recurrence-free survival; MFS, metastasis-free survival. Created using BioRender.

## Methods

### Study design and patient cohorts

The study cohort consisted of 102 formalin-fixed, paraffin-embedded (FFPE) tumor samples from 101 patients with a high-grade (grade 3) STS, including 51 patients with a UPS, 25 patients with a high-grade LMS, and 25 patients with a DDLPS. Clinical, pathologic, and treatment characteristics have been described previously [6].

The external validation cohort was composed by a group of 129 patients with comparable histopathological subtypes, which was selected from TCGA-SARC [8]. Cohort characteristics have also been described previously [6].

### Genomic alterations and sequence reconstruction

DNA and RNA sequencing (and the respective quality control) of the samples included in the study cohort has been already reported in detail in past works [6,9].

The genomic input consisted of reconstructable short variants, fusions and rearrangements. Short variants included single-nucleotide variants and insertions or deletions. Alteration records were converted into nucleotide sequence inputs representing the altered context. Fusions and other rearrangements contributed sequence inputs when sufficient information was available forreconstruction. The analysis therefore represented the sequence consequences of eligible events rather than a complete reconstruction of each tumor genome.

Short variants were reconstructed using Ensembl coding DNA sequences corresponding to GRCh37. Protein-coding transcripts were assessed in a predefined order prioritizing the canonical transcript, followed by longer coding sequences, and the first transcript compatible with the reported alteration was selected. Coding HGVS alterations were applied in transcript orientation to generate paired reference and alternative nucleotide inputs. The reconstruction procedure supported substitutions, deletions, insertions and deletion–insertions, checked coding-coordinate bounds and verified reference bases where explicitly specified.

RNA-reported fusion events were represented by breakpoint-independent proxy sequences, constructed by joining the terminal coding sequence of the first reported partner to the initial coding sequence of the second. Other rearrangements were not converted into sequence inputs. Copy-number alterations were retained as separate genomic annotations and were not encoded as Evo2 sequence features. Of 113 eligible short variants, 85 were successfully reconstructed, yielding 170 reference and alternative sequences. Together with the fusion proxies, these comprised 174 sequence inputs.

Regarding TCGA-SARC, somatic variant calls were obtained from the Genomic Data Commons masked somatic mutation files generated from whole-exome sequencing. Protein-altering short variants with available coding HGVS annotations and Ensembl transcript identifiers were selected. TCGA-SARC variants were reconstructed using Ensembl coding DNA sequences and the transcript specified in each mutation annotation. Unlike the GRCh37-based study-cohort pipeline, TCGA-SARC reconstruction used GRCh38 to match the reference assembly explicitly recorded in the GDC mutation files. The TCGA-SARC sequence analysis included short variants only, without fusion, rearrangement or copy-number sequence features. A total of 3649 were successfully reconstructed, yielding 7298 reference and alternative sequences.

In both cohorts, the analysis represented selected sequence consequences of eligible alterations rather than a complete reconstruction of each tumor genome.

### Evo2 feature extraction and patient representation

Reconstructed sequences were processed with Evo2 to obtain numerical representations of their sequence context [7]. Sequence-level features were then aggregated across alterations assigned to the same patient, yielding a patient-level genomic profile for survival modelling. This separates representation extraction from the subsequent modelling of clinical outcomes: Evo2 supplied genomic features, whereas the Cox models related those features to survival.

The pretrained evo2_7b (https://huggingface.co/arcinstitute/evo2_7b) was used with fixed model parameters, without cohort-specific or outcome-supervised fine-tuning. Sequences were evaluated in transcript orientation, without reverse-complement averaging. For each short variant, the reference and alternative sequences were scored independently using their mean autoregressive log-likelihood. The alternative-minus-reference difference (Δ) quantified the change in model-assigned sequence likelihood, consistent with the zero-shot likelihood-comparison principle illustrated in the NVIDIA BioNeMo Evo2 BRCA1 workflow (https://docs.nvidia.com/bionemo-framework/2.5/user-guide/examples/bionemo-evo2/zeroshot_brca1/). Variant scores were aggregated within patients to capture average and extreme sequence effects and thresholded disruption burdens.

Sequence embeddings were extracted from the *blocks.28.mlp.l3* layer and averaged across nucleotide positions. For each short variant, the Cosine distance between the pooled alternative and reference embeddings was calculated before standardization or dimensionality reduction. These displacements were summarised within each patient by their mean, maximum and sum. Additional covariates captured alternative-sequence likelihood and reconstructed short-variant burden. Aggregation was unweighted, without weighting by variant allele frequency, sequencing depth or gene identity.

For sequence-level dimensionality reduction, pooled sequence embeddings were standardised feature-wise and subjected to PCA separately within each cohort. Reference and alternative embeddings were included in both cohorts, together with fusion-proxy embeddings in the study cohort. Patient-level component summaries were then obtained by averaging the retained principal-component coordinates across each patient’s alternative short-variant sequences.

A complementary patient-level representation was constructed by averaging the original pooled alternative-sequence embeddings within each patient, including fusion-proxy embeddings where available and excluding reference embeddings. These patient-level vectors were standardized and subjected to a separate PCA fitted jointly across the study cohort and TCGA-SARC. This transformation therefore operated on patient-averaged original embeddings, rather than on the outputs of the sequence-level PCA.

Variant-score summaries, embedding-derived aggregates and patient-profile principal-component coordinates were concatenated to form the final Evo2 covariate vector. The constituent variables and preprocessing procedures are specified in *Supplementary Table 1*. Variant scores, embedding distances and burden terms were included without PCA; only embedding representations underwent dimensionality reduction. No further dimensionality reduction was applied to the concatenated vector.

In practical terms, each patient contributed one vector of numerical values to the Cox model. This vector combined mutation-score and burden summaries, the mean, maximum and total distance between reference and alternative embeddings, and the average alternative-sequence likelihood. It also included the patient-averaged sequence components PC1–PC3 and the patient-profile components PC1–PC5. Resulting this way on EVO2-derived genomic profiles.

### Comparator and integrated prognostic models

SARCULATOR represented the clinicopathologic comparator and CINSARC the transcriptomic comparator [3,4]. Cox proportional hazards models were used to evaluate Evo2-derived genomic profiles alone and in combination with each comparator. We use “SARCULATOR plus Evo2” and “CINSARC plus Evo2” to denote the corresponding integrated model families. These labels distinguish the source of the added information without implying that the underlying model specifications were identical between all endpoints.

Previously calculated SARCULATOR five-year overall-survival probabilities were imported from the clinical datasets. For the study cohort, these predictions were obtained using the site-specific extremity soft-tissue sarcoma or retroperitoneal sarcoma nomograms, as described previously. Regarding TCGA-SARC, the supplied five-year overall-survival predictions were used. In both cohorts, patients were classified according to predicted survival of ≤60% or >60%, with the latter serving as the reference category. This classification was retained across endpoints. The binary SARCULATOR-derived category, rather than the continuous survival probability or its individual clinicopathologic components, entered the Cox models.

CINSARC was evaluated in TCGA-SARC using the previously assigned C1 and C2 classifications supplied with the clinical data. Classifications were matched to patients using TCGA patient identifiers and entered as a binary covariate, with C1 as the reference category. Neither individual gene-expression measurements nor the accompanying continuous association score entered the survival models. Expression normalization, gene mapping and CINSARC classification were not repeated in the present analysis. CINSARC was not evaluated in the study cohort because its targeted RNA assay did not cover the complete signature, as previously reported.

Integrated models combined the complete Evo2 covariate vector described in *Supplementary Table 1* with the relevant comparator indicators as additive terms, without interactions. Let E denote the standardized Evo2 covariate vector, S the standardized indicator for SARCULATOR-predicted survival ≤60%, and C the standardized indicator for CINSARC class C2. The model-specific linear predictors were: Evo2: η = βLE; SARCULATOR plus Evo2: η = β□E + αS; CINSARC plus Evo2: η = β□E + γC; SARCULATOR plus CINSARC plus Evo2: η = β□E + αS + γC. For each model, the hazard was defined as h(t) = h₀(t) × exp(η), where h₀(t) denotes the unspecified baseline hazard. Coefficients were estimated independently for each model, cohort and endpoint; the notation does not imply shared coefficient estimates.

Models fitted in the present analysis used standardized predictors and ridge-penalised Cox regression, with Breslow handling of tied event times. The penalty parameter was 0.1, corresponding to subtraction of 0.05 times the sum of squared regression coefficients from the partial log-likelihood. The same Evo2 covariate definitions were retained across endpoints, although eligible populations and fitted coefficients differed. Reported C-indices from these models were apparent estimates calculated in the fitting population. Published comparator C-indices retained in the results tables were distinguished from estimates generated by these refitted models.

### Survival outcomes

In the study cohort, overall survival (OS) was measured from surgery to death from any cause. Disease-free survival (DFS) was measured from surgery to the first local recurrence, distant recurrence or death. Recurrence-free survival (RFS) was measured from surgery to the first local or distant recurrence; death without recurrence was treated as censoring, with observation ending at death or the last clinical assessment, whichever occurred first. Progression-free survival (PFS) was assessed from surgery to the earliest recorded recurrence, metastasis or death. For these endpoints, patients without an event were censored at their last recorded clinical assessment. Regarding TCGA-SARC, OS and metastasis-free survival (MFS) were assessed using the clinical durations and outcome annotations supplied with the previously published dataset. OS counted death from any cause, whereas MFS counted documented metastasis. Patients without the relevant event were censored at their recorded follow-up duration; death without metastasis was not counted as an MFS event [10].

### Model fitting and performance assessment

Prognostic discrimination was assessed using Harrell’s C-index [11]. Higher values indicate better ranking of observed survival outcomes among comparable patient pairs. For each comparator, the absolute difference in C-index was calculated as the integrated-model estimate minus the comparator-only estimate. We report these differences on the original C-index scale. They are not relative improvements in survival or estimates of treatment benefit.

The principal comparisons were SARCULATOR versus SARCULATOR plus Evo2 within each cohort and CINSARC versus CINSARC plus Evo2 within TCGA-SARC. Values are reported to three decimal places. We interpret the comparisons descriptively and do not infer statistical significance from point estimates alone. The C-index can depend on censoring and case mix; comparisons across cohorts therefore have a different interpretation from comparisons between models evaluated in the same patients [12].

### Ethics and reproducibility

The present study was performed in accordance with the ethical standards of Helsinki Declaration II and was approved by the Institution Review Boards of both Centro Académico de Medicina de Lisboa and Instituto Português de Oncologia de Lisboa Francisco Gentil.

## Results

### Cohort composition and genomic representation

The study cohort included 102 patients: 25 with DDLPS (24.5%), 25 with high-grade LMS (24.5%) and 52 with UPS (51.0%). The separate TCGA-SARC cohort comprised 129 patients with comparable histologies. Evo2 representations were generated from 174 sequences in the study cohort and 7298 in TCGA-SARC. These counts describe sequence inputs and do not replace patient-level analytical denominators.

The workflow linked individual alteration sequences to a shared patient representation before survival modelling (Figure 1). This enabled genomic features to be evaluated with either a clinicopathologic comparator or a transcriptomic comparator. The numbers of reconstructed sequences differed substantially between cohorts, making the coverage and composition of the genomic inputs relevant to subsequent interpretation.

### Integration with SARCULATOR in the study cohort

Adding Evo2-derived features to SARCULATOR increased the reported C-index for each of the four study-cohort endpoints (Table 1; Figure 2a). For OS, the C-index increased from 0.620 to 0.770, an absolute difference of 0.150. For PFS, the corresponding values were 0.625 and 0.755, a difference of 0.130. For DFS, the C-index increased from 0.612 to 0.714, a difference of 0.102. For RFS, it increased from 0.633 to 0.771, a difference of 0.138.

**Figure 2.**
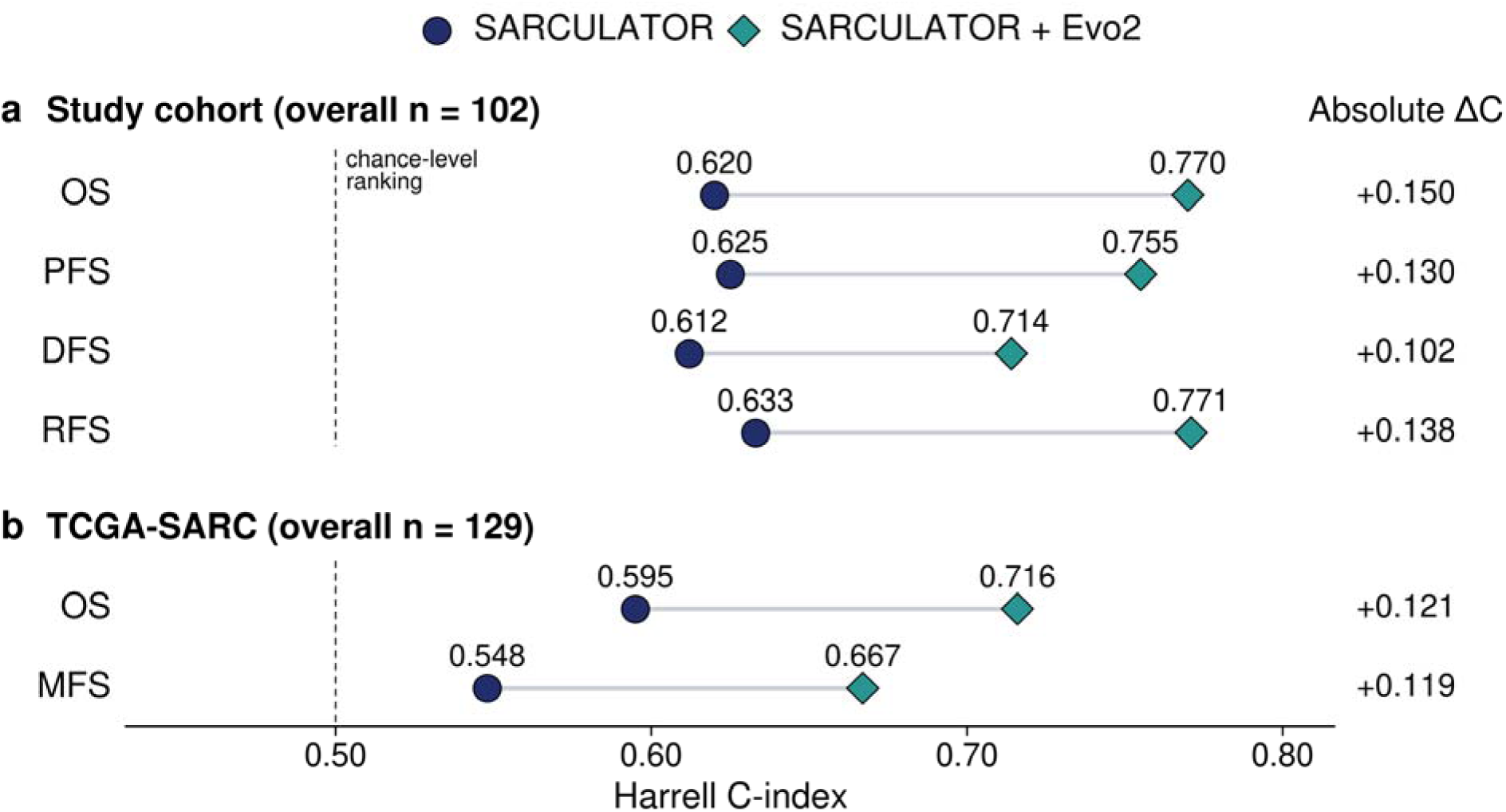
**Integration of Evo2 genomic features with SARCULATOR**. Harrell’s concordance index (C-index) for SARCULATOR alone and SARCULATOR plus Evo2 in **a**, the study cohort and **b**, TCGA-SARC. Navy circles indicate the clinicopathologic comparator and teal diamonds the integrated model. Grey connecting segments join corresponding point estimates and do not denote confidence intervals. Absolute ΔC is the integrated-model C-index minus the comparator C-index. The dashed vertical line marks C-index = 0.5. Comparisons are descriptive and do not establish statistical significance. Displayed sample sizes are overall cohort sizes, not endpoint-specific analytical denominators. OS, overall survival; PFS, progression-free survival; DFS, disease-free survival; RFS, recurrence-free survival; MFS, metastasis-free survival; TCGA-SARC, The Cancer Genome Atlas sarcoma cohort.

**Table 1.** Discrimination of comparator and integrated models.

| Cohort | Endpoint | Comparator | C-index<br>Comparator | C-index<br>+ Evo2 | Absolute<br>$\Delta C$ |
| --- | --- | --- | --- | --- | --- |
| Study cohort | OS | SARCULATOR | 0.620 | 0.770 | +0.15 |
| Study cohort | PFS | SARCULATOR | 0.625 | 0.755 | +0.130 |
| Study cohort | DFS | SARCULATOR | 0.612 | 0.714 | +0.102 |
| Study cohort | RFS | SARCULATOR | 0.633 | 0.771 | +0.138 |
| TCGA-SARC | OS | SARCULATOR | 0.595 | 0.716 | +0.121 |
| TCGA-SARC | MFS | SARCULATOR | 0.548 | 0.667 | +0.119 |
| TCGA-SARC | OS | CINSARC | 0.476 | 0.653 | +0.177 |
| TCGA-SARC | MFS | CINSARC | 0.553 | 0.650 | +0.097 |

**Discrimination of comparator and integrated models.** The integrated model adds Evo2-derived genomic features to the named comparator. Absolute $\Delta C$ = C-index of the integrated model minus C-index of the comparator. All values are descriptive point estimates. They are not relative improvements in survival. Confidence intervals and endpoint-specific sample and event counts remain to be completed.
OS, overall survival; PFS, progression-free survival; DFS, disease-free survival; RFS, recurrence-free survival; MFS, metastasis-free survival. No cross-cohort pooling was performed for this table.

The integrated model estimates ranged from 0.714 to 0.771. The direction of the difference was consistent across OS and the three non-OS endpoints. These endpoints are related and may include overlapping events; their concordant direction should therefore be interpreted as within-cohort consistency rather than four independent replications.

### Integration with SARCULATOR in TCGA SARC

In TCGA-SARC, the OS C-index was 0.595 for SARCULATOR and 0.716 for SARCULATOR plus Evo2, an absolute difference of 0.121. For MFS, the corresponding estimates were 0.548 and 0.667, a difference of 0.119 (Table 1; Figure 2b).

The direction of the OS comparison agreed with that observed in the study cohort. The additional MFS comparison supported evaluation of the genomic representation for metastatic outcomes. Differences in cohort composition, available genomic inputs and endpoint ascertainment preclude attributing between-cohort variation in C-index to the representation alone.

### Integration with CINSARC in TCGA SARC

CINSARC annotation of the study cohort was not possible since the FoundationOne RNA gene set that was used does not include 32 of the genes included in CINSARC (48% of the total number of genes considered in CINSARC). CINSARC annotation of a cohort requires that all of the 67 genes that compose this molecular signature are covered by the gene set of the sequencing test that is employed to map the transcriptomic landscape of that cohort.

In TCGA-SARC, adding Evo2-derived features to CINSARC increased the reported OS C-index from 0.476 to 0.653, an absolute difference of 0.177. The MFS C-index increased from 0.553 to 0.650, a difference of 0.097 (Table 1; Figure 3).

**Figure 3.**
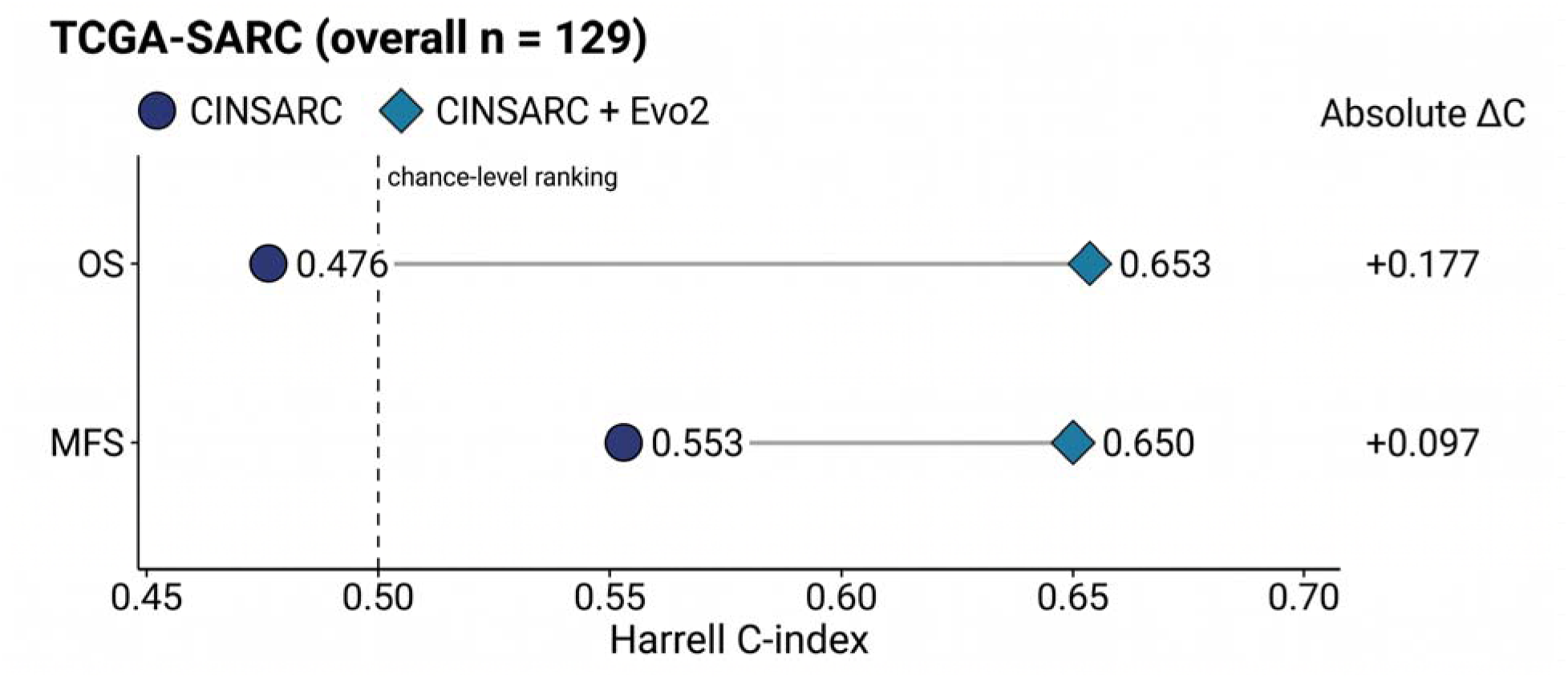
Integration of Evo2 genomic features with CINSARC. Harrell’s concordance index (C-index) for CINSARC alone and CINSARC plus Evo2 in TCGA-SARC. Navy circles indicate the transcriptomic comparator and blue diamonds the integrated model. Absolute ΔC is the integrated-model C-index minus the comparator C-index. Grey connecting segments join corresponding point estimates and do not denote confidence intervals. The dashed vertical line marks C-index = 0.5. Comparisons are descriptive and do not establish statistical significance. The overall cohort size does not specify endpoint-specific analytical denominators. Estimates describe the evaluated CINSARC implementation, not its performance in other settings. OS, overall survival; MFS, metastasis-free survival; TCGA-SARC, The Cancer Genome Atlas sarcoma cohort.

These comparisons extend the integration analysis to a transcriptomic predictor. The OS C-index for CINSARC alone was below 0.5. The results describe the evaluated CINSARC implementation in this cohort and should not be taken as a general estimate of the signature’s performance in its original setting.

### Evo2 alone compared with SARCULATOR and CINSARC

In the study cohort, Evo2 alone yielded higher reported C-index estimates than SARCULATOR across all four endpoints. For OS, the C-index was 0.703 for Evo2 versus 0.620 for SARCULATOR, an absolute difference of 0.083. Corresponding estimates were 0.736 versus 0.625 for PFS, 0.644 versus 0.612 for DFS, and 0.676 versus 0.633 for RFS, representing absolute differences of 0.111, 0.032 and 0.043, respectively.

Regarding TCGA-SARC, Evo2 alone also yielded higher reported discrimination than SARCULATOR. For OS, the C-index was 0.657 versus 0.600, an absolute difference of 0.057. For MFS, the corresponding estimates were 0.638 versus 0.548, a difference of 0.090.

Evo2 alone likewise yielded higher reported C-index estimates than CINSARC in TCGA-SARC. For OS, the C-index was 0.657 for Evo2 versus 0.490 for CINSARC, an absolute difference of 0.167. For MFS, the corresponding estimates were 0.638 versus 0.553, a difference of 0.085.

These comparisons describe apparent Evo2 performance relative to the reported comparator estimates. Because published comparator benchmarks were retained where applicable, the differences should not be interpreted as paired estimates establishing statistical superiority or externally validated performance.

### Overall pattern of model comparisons

Across the eight comparator-versus-integrated-model comparisons, all reported C-index differences were positive, ranging from 0.097 to 0.171. Adding Evo2-derived features to SARCULATOR or CINSARC therefore consistently increased point-estimate discrimination across the evaluated cohorts and endpoints.

Evo2 alone also yielded higher reported C-index estimates than SARCULATOR across the four study-cohort endpoints: OS, 0.703 versus 0.620; PFS, 0.736 versus 0.625; DFS, 0.644 versus 0.612; and RFS, 0.676 versus 0.633. The corresponding absolute differences ranged from 0.032 to 0.111.

Regarding TCGA-SARC, Evo2 alone achieved an OS C-index of 0.657, compared with 0.600 for SARCULATOR and 0.490 for CINSARC. For MFS, the corresponding estimates were 0.638, 0.548 and 0.553, respectively. Thus, Evo2-only estimates exceeded the reported SARCULATOR estimates by 0.057 for OS and 0.090 for MFS, and the CINSARC estimates by 0.167 and 0.085.

Together, these results show numerically higher reported discrimination for Evo2 alone and for models integrating Evo2 with established comparators. These comparisons remain descriptive: the Evo2 estimates are apparent, published comparator benchmarks were retained where applicable, and paired confidence intervals are unavailable. The findings do not establish statistical superiority, externally validated performance or a causal mechanism of tumour aggressiveness.

## Discussion

This study shows that integrating Evo2-derived representations of altered tumor sequences with established sarcoma prognostication models increased reported prognostic discrimination across two cohorts. All eight comparator-versus-integrated-model comparisons favored the addition of genomic features, with absolute C-index gains of 0.097–0.171. For OS, SARCULATOR plus Evo2 increased the C-index from 0.620 to 0.770 in the study cohort and from 0.595 to 0.716 in TCGA-SARC; CINSARC plus Evo2 increased it from 0.476 to 0.653 in TCGA-SARC. These observations are consistent with genomic foundation models contributing a substantial, partly independent prognostic signal beyond clinicopathologic and transcriptomic risk stratification systems. The improvement after integration suggests information not fully captured by either comparator, although formal statistical independence requires appropriate adjustment and reliable out-of-sample evaluation.

Previous studies have demonstrated that protein-language-model-or AlphaFold-derived variant-effect estimates can stratify clinical outcomes, predominantly in lung cancer or treatment-selected pan-cancer cohorts. However, these investigations have generally been restricted to protein-coding missense variants, selected cancer genes or categorical pathogenicity assignments. To our knowledge, no previous study has evaluated an aggregated, nucleotide-level genomic foundation model effect score as an independent prognostic signal in soft tissue sarcoma or examined its incremental value beyond both established clinicopathologic and transcriptomic risk models.

The central contribution is the connection between alteration-level sequence information and patient-level survival modelling. A sequence representation retains information that is not explicitly encoded by a clinicopathologic score or a fixed gene expression signature. For a short variant, that information may include the surrounding nucleotide context. For a fusion or rearrangement, it may include the newly formed sequence junction. Aggregating such representations allows the combined set of reconstructable alterations to be analyzed at the patient level. The resulting profile therefore provides a way to relate the sequence composition of an individual tumor to its clinical trajectory without requiring a predefined prognostic gene signature.

This distinction has biological relevance because the meaning of a genomic alteration depends on its context. Changes affecting coding sequence, splice-associated elements or regulatory motifs may have different consequences even when assigned to the same gene or alteration category. Evo2’s published analyses show that its representations capture features associated with evolutionary constraint, exon–intron organization and regulatory sequences [7]. Such capabilities provide a rationale for exploring whether the reconstructed tumor sequences contain information associated with malignant behavior. They do not imply that the present embeddings measure the functional effects of every alteration. Because altered-sequence embeddings can also encode the identity and composition of the underlying locus, the contribution of the alteration itself must be distinguished from that of its sequence background.

A shared representation space may be especially useful in rare, heterogeneous cancers, where individual alterations are less frequent and gene-by-gene encodings become sparse. Pretraining could allow related sequence features to contribute across different events, reducing dependence on observing the same alteration repeatedly in a small clinical cohort. Biologically, this could help detect convergent disruption of processes governing genome maintenance, cellular differentiation or adaptation despite distinct mutational events. This remains a hypothesis for the observed prognostic gains. Neither survival discrimination nor attribution to an embedding establishes a sarcoma-specific mechanism, and technical dissimilarities or differential subtype composition may also contribute to the signal. Comparisons with simpler genomic encodings and independent functional evidence are therefore essential.

The complementarity with SARCULATOR is conceptually straightforward. Clinicopathologic nomograms combine accessible, clinically interpretable features that summarize tumor burden, morphology and patient characteristics and have undergone substantial external validation [3]. However, patients with similar nomogram predictions may harbor different biological determinants of progression. Sequence-derived features could resolve some of this residual heterogeneity by representing molecular variation within otherwise similar clinical categories. Conversely, clinical variables retain information about the patient and disease setting that a tumor sequence cannot provide. The higher discrimination of SARCULATOR plus Evo2 is therefore compatible with integration of two partially overlapping sources of information, rather than evidence that clinical assessment has become dispensable.

The relationship with CINSARC addresses a different limitation. CINSARC captures a validated transcriptional programme centered on mitosis and chromosome maintenance [4], whereas sequence representations interrogate the genomic contexts of eligible alterations. A finite expression signature does not capture all perturbations associated with aggressiveness, especially those outside its selected gene coverage or those whose effects vary with the altered sequence. Genomic features could consequently add information beyond the measured programme without assuming that they are universally more informative than transcriptional state. There is already precedent for integrating molecular and clinical risk: combining CINSARC with SARCULATOR improved prognostic discrimination in a separate retrospective sarcoma study [13]. The present findings extend this principle to learned representations of altered DNA sequences.

Our previous transcriptomic classification provides a further biological context. Its four transcriptomic subtypes reflected variation in DNA-repair gene expression, cancer-testis antigens, MHC class II/antigen-presentation gene expression and claudin-associated programmes across conventional histological categories [6]. These observations suggest that clinically relevant diversity extends beyond a single proliferative programme or morphological label. A subsequent joint analysis of SARCULATOR, CINSARC, these transcriptomic subtypes and Evo2-derived features could determine which information is shared and which remains complementary. The current study does not establish superiority over the transcriptomic classification. More broadly, integrated models outperforming their respective comparators alone should not be conflated with universal superiority of Evo2 as a standalone predictor; the latter requires matched comparisons using identical patients, endpoints and evaluation procedures.

Comparator performance must also be interpreted within its intended setting. SARCULATOR includes models developed for specific anatomical and clinical populations, while CINSARC performance depends on endpoint, specimen characteristics and implementation and has varied in chemotherapy-treated cohorts [3–5]. In the present TCGA analysis, the below-chance OS point estimate for CINSARC warrants scrutiny of gene coverage, expression processing and risk-direction coding before the magnitude of the improvement is generalized. Moreover, incomplete coverage of CINSARC genes in the study-cohort assay restricted transcriptomic comparisons to TCGA-SARC. These considerations reinforce the importance of evaluating incremental value against faithfully implemented comparators rather than attributing every difference to the genomic representation.

The use of a separate TCGA-SARC cohort broadens the empirical evaluation beyond the study cohort, but independent patient origin and external model validation are different properties. External validation of a prediction model requires the fitted prediction rule and its preprocessing to be established before application to the evaluation cohort. If coefficients, feature selection or tuning were re-estimated using TCGA-SARC outcomes, the analysis would instead test a modelling approach in a second dataset. Clear separation of these designs is essential to understanding what the results show [11,14].

The large difference in sequence-input counts also requires attention. It could reflect sequencing coverage, available alteration classes, reconstruction rules or other differences between datasets. It does not demonstrate a biological difference in mutation burden. A representation aggregated across many sequences may behave differently from one based on only a few sequences. Sensitivity analyses restricted to comparable genomic coverage and event classes, together with assessment of the number of sequences per patient, would help establish whether the prognostic association persists under more comparable inputs.

Pretraining provenance is another aspect of transportability. The exact checkpoint and its documented training sources should be recorded. Exposure to a reference sequence during self-supervised pretraining is not equivalent to exposure to a patient’s survival outcome. Nevertheless, overlap with evaluation sequences, if present, should be described rather than assumed absent. The clinical evaluation must independently ensure that outcome information does not influence preprocessing, representation selection or model tuning in the test cohort.

The potential clinical impact lies in reducing uncertainty at decisions for which current risk estimates leave several reasonable options. In localized STS, better calibrated estimates could refine multidisciplinary discussions about the employment of perioperative systemic therapy conceptually, the type of perioperative systemic therapy to use, trial enrolment and surveillance strategies, particularly for patients near a clinically relevant risk threshold. Retrospective analysis of EORTC-STBSG 62931 illustrates why individual risk matters: the benefit associated with adjuvant chemotherapy was concentrated in patients with a poor SARCULATOR-predicted prognosis [2]. Nevertheless, the threshold and treatment implications of an established model cannot simply be transferred to a new score. It is important to underline that a patient reclassified as high risk by Evo2 would not automatically be more chemotherapy-sensitive and more prone to respond to different chemotherapy regimens. Decisions on the use of radiotherapy and different types of surgical approaches also depend on individual clinical characteristics, such as local anatomy, resectability and local-control risk, that have to be interpreted and integrated on a case-to-case basis and which a survival score cannot replace [1].

Clinical usefulness therefore requires more than a higher C-index. Discrimination measures how well a model ranks outcomes; it does not establish the accuracy of an individual absolute risk estimate and does not quantify a gain in survival [11,12]. Calibration at prespecified time horizons and decision-curve analysis should determine whether the additional information improves decisions at relevant thresholds [15]. Prospective evaluation should measure whether model-informed care reduces avoidable toxicity, overtreatment or undertreatment, improves quality of life and outcomes, and remains feasible in routine therapeutic pathways. Turnaround time, assay failure, tissue requirements, computational cost and access to sequencing could materially affect clinical value. Any proposal to intensify surveillance or safely reduce treatment requires evidence of benefit in that specific use case.

The statistical interpretation remains limited by retrospective sampling, modest cohort sizes, unreported endpoint-specific event counts and the complexity of the genomic representation. The effective sample size is determined by independent patients and outcome events, not by the number of reconstructed sequences [11,16]. Feature selection, normalization, dimensionality reduction and model tuning must therefore be included within the training and validation procedure, with patient-level separation of all samples and sequences. Paired confidence intervals for C-index differences, optimism correction and assessment of proportional-hazards assumptions are needed. Related survival endpoints can share events and should not be counted as independent replications. Differences in follow-up, censoring and endpoint definitions also require attention, particularly when deriving MFS from TCGA clinical fields [10,12].

Biological coverage imposes additional limitations. The present representation includes alterations that could be reconstructed as sequence inputs and consequently may omit relevant events or patients with insufficient information. Copy-number changes are prominent in adult STS [8], but nucleotide sequence representations do not automatically encode gene dosage, clonality or the complete chromosomal architecture. Similarly, they do not directly measure transcription, post-translational/transcriptional modifications, protein activity, microenvironmental immune-cell composition or metabolic flux patterns in and between different cell and microenvironmental compartments. Our observation of SDHB overexpression associated with enzymatic dysfunction in UPS illustrates why a molecular abundance signal cannot always be equated with its functional consequence [9]. These considerations favor concomitantly testing complementary molecular measurements rather than assuming that a single representation captures the full biology of the tumor.

The immediate next step is evaluation of a fully specified, locked model in geographically and temporally independent cohorts, followed by prospective assessment in its intended clinical population. Comparisons should include alteration burden, gene-level indicators, explicit copy-number features, conventional sequence summaries and alternative pretrained representations. Reference-versus-altered sequence analyses, removal of selected event classes and stratification by subtype, anatomical site and type of treatment exposure could clarify the source and stability of the signal. Attribution to individual sequences or positions should be reproducible across model fits and linked to independent experimental observations. Joint assessment with RNA, protein, metabolite and functional measurements would help distinguish an interpretable biological association from a technically useful but mechanistically unresolved predictor.

A broader opportunity is to develop foundation-model-guided stratification that addresses both prognosis and therapeutic vulnerability. This would require separate, explicitly evaluated objectives: estimating outcome under a defined clinical setting and identifying differential benefit from a particular intervention. Evo2 representations could help prioritize candidate alterations or molecular programmes for functional testing and support treatment-specific models trained with drug-response or clinical-treatment data. However, sequence plausibility, predicted functional disruption and clinical actionability are distinct concepts. Treatment approaches should be interpreted through a molecular tumor board and graded against clinical evidence, for example using the ESMO Scale for Clinical Actionability of molecular targets [17]. The current survival models do not establish a treatment-response biomarker.

DNA-repair vulnerabilities provide one concrete avenue. Sequence features suggesting disruption of repair-associated loci could be investigated alongside transcriptomic programmes, genomic scars and functional repair assays to identify candidates for DNA-damaging or synthetic-lethal strategies. A single-arm phase II study of olaparib plus temozolomide in advanced uterine leiomyosarcoma reported encouraging activity and incorporated functional assessment of homologous recombination [18]. However, the subsequent biomarker-unselected randomized Alliance A092104 trial did not demonstrate a progression-free survival advantage, as reported in its conference abstract [19]. This contrast supports the need for more discriminating biomarker hypotheses; it does not establish that an Evo2 score identifies responsive patients. Findings in uterine LMS also cannot be generalized to all LMS or other sarcomas. Foundation-model prioritization would therefore need to be followed by confirmation of the relevant repair defect and prospective evaluation of treatment benefit, including resistance and toxicity.

Other modalities require different molecular evidence. For a candidate oncogenic kinase fusion, sequence-based prioritization could be combined with confirmation of an expressed, functional target; the clinical activity of larotrectinib in TRK fusion-positive cancers illustrates an established route from a specific molecular lesion to matched inhibition [20]. This example applies only when an actionable fusion is verified and should not be generalized to all rearrangements. For immune therapies, integrating genomic features with antigen expression, HLA genotype and tumor immune context could generate hypotheses about checkpoint blockade or antigen-directed cell-based immunotherapy. The SU2C-SARC032 trial of perioperative pembrolizumab in selected high-risk extremity sarcomas provides a clinically relevant exemplificative setting for future predictive biomarker studies [21]. The MAGE-A4-directed T-cell therapy evaluated in SPEARHEAD-1 provides a separate example of antigen-and HLA-dependent treatment selection in synovial sarcoma and myxoid round-cell liposarcoma [22]. The above-mentioned histotypes were not included here, and prognostic embeddings alone cannot establish antigen expression, antigen presentation or treatment eligibility.

These therapeutic directions could be investigated using foundation-model-prioritized hypotheses followed by perturbation studies and drug testing in models that retain relevant tumor and microenvironmental features. The metabolic phenotype identified in our UPS work additionally supports exploration of experimentally confirmed metabolic dependencies [9]. Responses in such systems would remain preclinical evidence, but could guide molecularly stratified trials of cytotoxic, targeted, immune or combination approaches. Clinical evaluation should test treatment-by-biomarker interactions wherever possible, because poor prognosis alone cannot distinguish a sensitive tumor from an aggressive but resistant one. Ultimately, a useful framework would report both a validated risk estimate and evidence-graded therapeutic hypotheses, with uncertainty made explicit. The present findings provide a rationale for pursuing that integration; they establish a prognostic research direction rather than a treatment-selection system.

## Conclusions

Evo2-derived representations of altered tumour sequences were associated with higher reported C-index estimates when integrated with SARCULATOR or CINSARC in two sarcoma cohorts. These findings support the further study of genomic sequence representations as complementary prognostic biomarkers. Validation of a fixed prediction rule, together with assessment of calibration and clinical utility, is necessary to establish the reliability and clinical relevance of this approach.

## Declarations

### Data and code availability

TCGA-SARC is a publicly described research resource [1]. Somatic mutation data were obtained from the NCI Genomic Data Commons (https://portal.gdc.cancer.gov/projects/TCGA-SARC) under project identifier TCGA-SARC, using open-access whole-exome masked somatic mutation files. Data and Code will be made available upon publication.

## Data Availability

All data produced in the present study are available upon reasonable request to the authors.

## Acknowledgements

The authors would like to thank F.Hoffmann-LaRoche AG and Foundation Medicine Inc. for their support in the study cohort genomic and transcriptomic analyses, namely for providing the FoundationOne^®^CDx and FoundationOne^®^RNA assays, and for all the technical support, specifically during the transfer of the sequencing data via the safe platform. Moreover, the authors acknowledge the patients and research teams who contributed to TCGA.

## Use of artificial intelligence in manuscript preparation

ChatGPT 6 Astra was used to assist with drafting, literature checking and figure preparation, and BioRender was used to generate schematic figure drafts.

---

**Supplementary Table 1:**
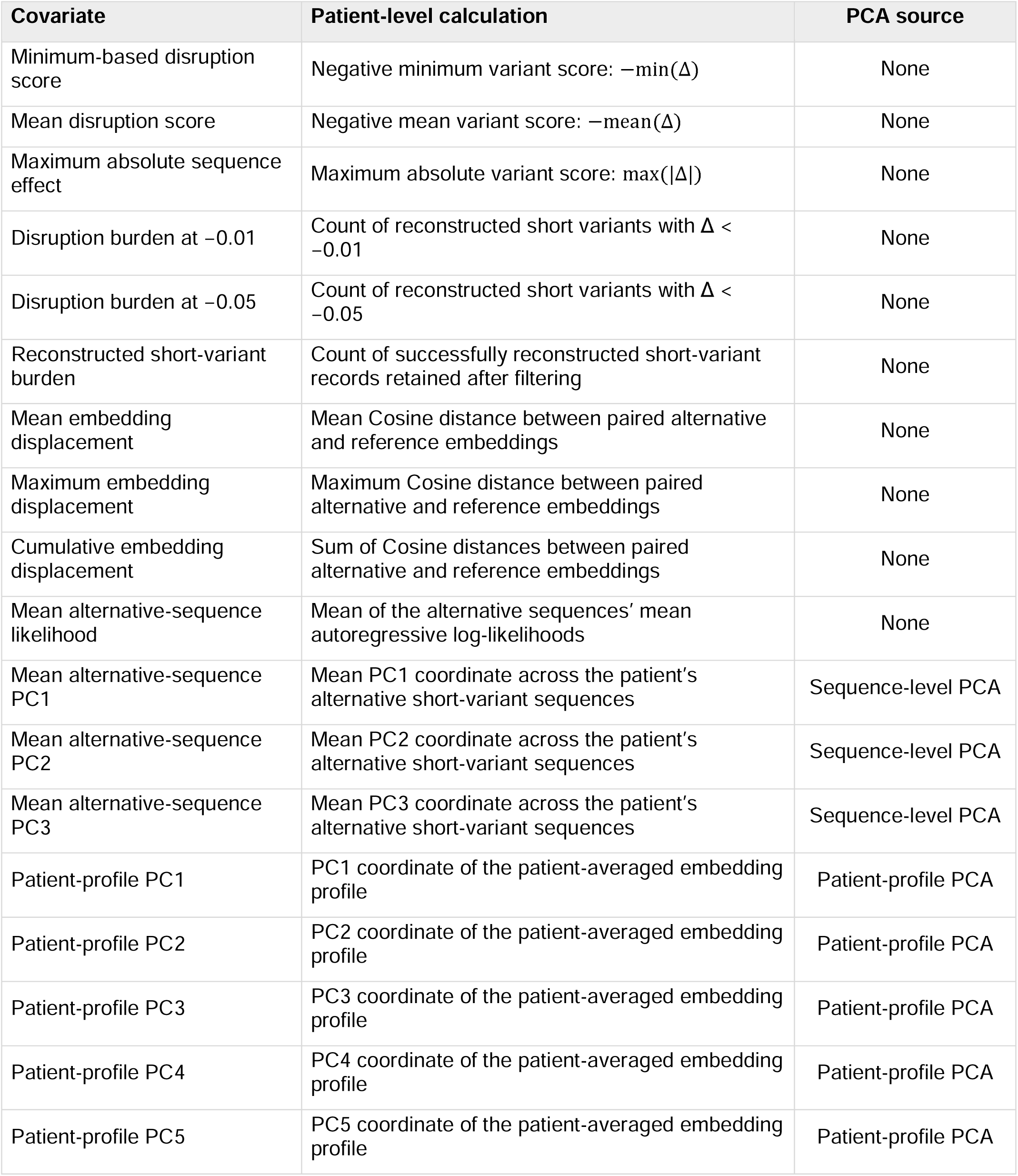
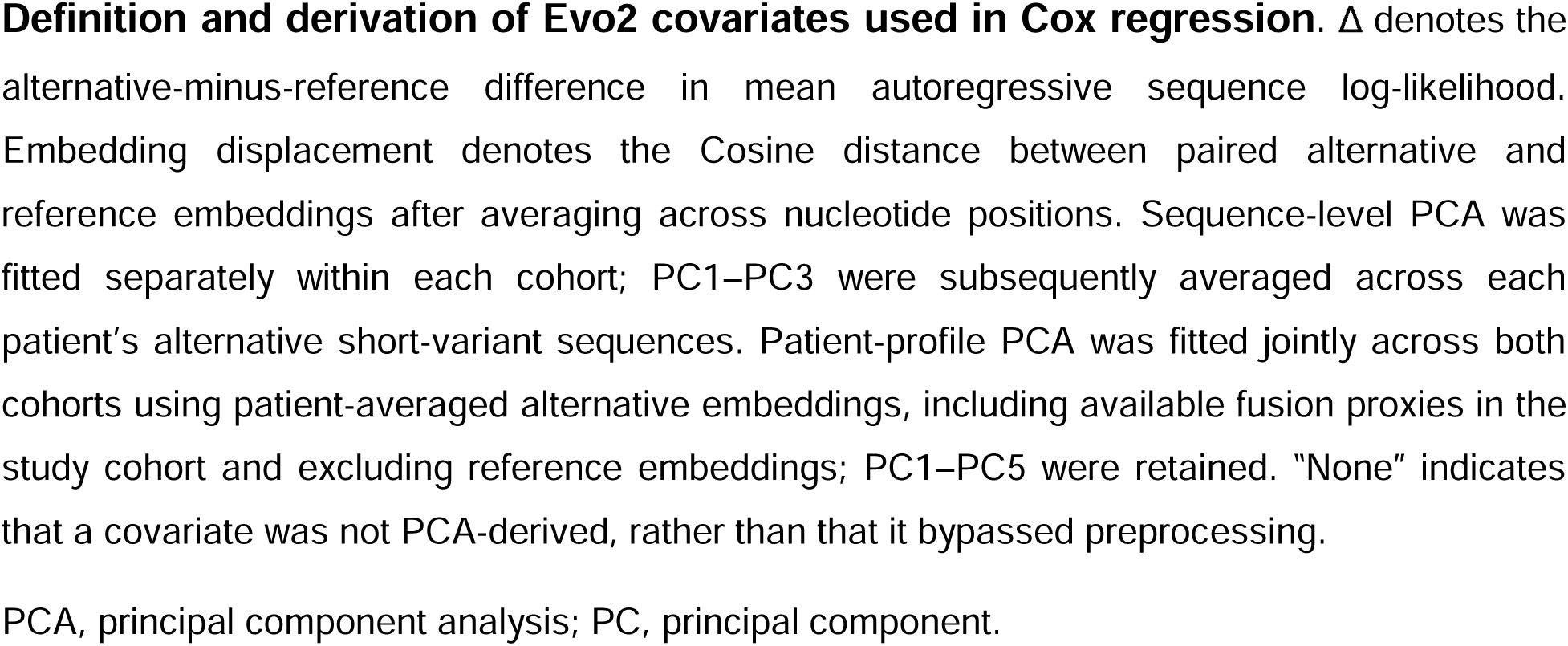
Definition and derivation of Evo2 covariates used in Cox regression.

